# Vaginal Microbiome and Preterm Birth in Pregnant Indian Women

**DOI:** 10.64898/2026.02.19.26346663

**Authors:** Abhishek Singh, Karisma Chhabria, Neerja Vashist, Salomi Singh, Garima Suneja, Shradha Khater, Amrita Rao, Chandrasekar Arigela, Akib Kamani, Gautam Das, Atahar Husein, A.D Urehekar, Sushil Kumar, Deepak Modi

**Author notes:** Corresponding author Dr. Abhishek Singh, Assistant professor, Department of Microbiology, NC Medical College, Israna Haryana India. E.mail.

## Abstract

**Objective:** Preterm birth (PTB) is a leading cause of neonatal morbidity and mortality worldwide, with India alone contributing nearly 27% of the global PTB burden. Although alterations in the vaginal microbiome have been implicated in PTB, its association in the Indian context is underexplored. This study aimed to investigate the association of vaginal microbiome and PTB in Indian women at the time of delivery.

**Study design:** The vaginal swabs were collected at the time of delivery from 72 women (31 term, 41 preterm) admitted to a tertiary care hospital in Western India. Microbial DNA was extracted, and the V3–V4 region of the 16S rRNA gene was sequenced. Community composition, alpha and beta diversity, and differential taxonomic abundance were assessed using bioinformatics pipelines.

**Results:** At the time of delivery, there were no significant differences in alpha or beta diversity between term and preterm groups. Principal coordinate and unsupervised clustering analyses showed no group-wise segregation. The relative abundance of individual *Lactobacillus* species, including *L. iners* and *L. helveticus*, did not differ significantly between the two groups. However, a modest difference in the relative abundance of *Streptococcus* was observed between the two groups after adjustment.

**Conclusion:** This study found no major microbial shifts in the vaginal microbiome associated with preterm birth in this cross sectional cohort of Indian women, suggesting that vaginal dysbiosis at the time of delivery may not be a principal driver of PTB in this population. These findings underscore the need for larger, longitudinal, and ethnically diverse studies using standardized methodologies better to understand the microbiome’s role in PTB risk.

## 1. Introduction

Preterm birth (PTB), defined as birth occurring before 37 completed weeks of gestation, constitutes a significant global health challenge. It is one of the leading causes of neonatal mortality and morbidity in both developed and developing nations, contributing to approximately 40% of neonatal deaths worldwide. Roughly 10% of pregnancies result in preterm birth globally(1), with approximately 3.6 of the 27 million annual births in India occurring preterm (2,3). Thus India contributes approximately 27% of the world’s preterm births making India the largest single-country contributor to the global PTB burden(1). This, highlights an urgent public health challenge and a key priority for the Indian government. Despite its clinical significance, the underlying mechanisms leading to PTB remain incompletely understood.

In recent years, evidence strongly suggests a link between vaginal microbial communities and the occurrence of preterm PTB. In healthy, non-pregnant women of reproductive age, the vaginal microbiome is usually less diverse and typically exhibits a predominance of *Lactobacillus* species, with most women displaying a prevalence of one specific species among *L. crispatus*, *L. iners*, *L. jensenii*, *and L. gasseri* (4–7). During pregnancy, the vaginal microbiome stabilizes further, characterized by a high abundance of *Lactobacillus* species(4,5,8,9). The *Lactobacillus*-dominated ecosystem fosters the maintenance of vaginal homeostasis and acts as a barrier against the colonization and proliferation of harmful microorganisms, including those associated with sexually transmitted infections (10,11). These protective mechanisms are multifaceted, encompassing vaginal pH reduction, production of bioactive compounds, competition for essential nutrients and attachment sites, and the modulation of the host’s immune response (10,12–14).

Several studies, predominantly from Western and East Asian populations, have reported associations between reduced Lactobacillus abundance and/or increased presence of anaerobic bacteria and the risk of preterm delivery. Pathogenic bacteria such as *Streptococcus agalactiae* (Group B *Streptococcus* or GBS), *Gardnerella vaginalis*, *Aerococcus*, *Prevotella bivia*, *Atopobium*, and *Clostridium* have been observed at higher relative abundances in women who experience PTB(15–19). Besides, the overall composition of the vaginal microbiome beyond the presence of individual bacterial species has also been implicated in various pregnancy complications, including infertility, abortions, stillbirths, and PTB (4,20–24). While several studies have demonstrated a significant association between disrupted microbiome and PTB but others have failed to confirm these findings and found no association of PTB and vaginal dysbiosis (24–30). Such discrepancies may be attributed to methodological differences, including sampling time points, sequencing platforms, and bioinformatics pipelines, as well as host-specific factors such as ethnicity, diet, hygiene practices, and regional antimicrobial exposures (24,30,31).

To date, most of the available data on the vaginal microbiome and its association with preterm birth focuses on populations of European descent, including American Whites, Blacks, Hispanics, and Europeans (25,26,28,29,31,32). Data from Asian populations is largely dominated by studies conducted on Chinese cohorts (33). Limited information is available regarding the vaginal microbiota and preterm birth in the Indian context. Three studies from India reported higher abundance of *Lactobacillus iners*, *Megasphaera* spp., *Gardnerella vaginalis*, and *Sneathia sanguinegens* in women who delivered preterm, the studies have a relatively small sample size or are from a single cohort (27,34–36). Given the lack of extensive data, further research on link of maternal vaginal microbiome and preterm birth in the Indian context is necessary.

In this study, we investigated the vaginal microbiome at the time of delivery in a cohort of Indian women with term and preterm births using 16S rRNA gene sequencing. Our goal was to determine whether microbial diversity or specific taxa are associated with PTB at the time of birth in this underrepresented population.

## 2. Materials and Methods

### 2.1 Ethical Approval

This study was approved by the Human Ethics Committee of MGM Institute of Health Sciences (MGMIHS), Kamothe, Navi Mumbai (Approval No: MGMIHS/RES/02/2020-21/401A). Before sample collection, written informed consent was obtained from all participants.

### 2.2 Study Design and Subjects

This is a cross-sectional case-control study. Pregnant women visiting the antenatal clinic at MGM Hospital Kalamboli were included in the study. Inclusion criteria were singleton pregnancies with no known history of gynaecological or obstetrical complications. Participants presenting with signs of labour (mild to strong contractions lasting at least one hour) at the hospital and delivered before completing 37 weeks of gestation were included in PTB group, while those presenting with signs of labour and delivering after 38 weeks of gestation were included in term group. Only those participants who had undergone at least two antenatal ultrasound examinations, with an estimated gestational age that aligned with the calculation based on the Last Menstrual Period (LMP), were included. Women whose LMP-based and ultrasound-based gestational ages differed by more than 1.5 weeks were excluded. Additionally, women who presented with obstetric complications such premature rupture of membranes, gestational diabetes, preeclampsia, hypertension, intrauterine growth restriction (IUGR), a history of fever, or symptoms of urinary tract infection were excluded from the study. Women on antibiotic or probiotic during previous 8 weeks in pregnancy were also excluded.

### 2.3 Collection of high vaginal swabs

High vaginal swabs were aseptically samples from the participants by a trained staff. The swabs were collected from the midpoint of the vagina using sterile cotton swab collectors (Hi-media) and placed in sterile glass tubes. The tubes were immediately sealed and stored at 4°C until transported to the laboratory within 60 minutes. A total of 72 samples that included 31 full-term deliveries (>38 weeks of gestation) and 41 preterm deliveries (<37 weeks of gestation) were subjected to microbiome analysis.

### 2.4 Extraction of Microbial DNA from vaginal swabs

Microbial genomic DNA was extracted from the high vaginal swab samples using the FavorPrep Soil DNA Isolation Kit (Cat. No. FAS01001-1, Favorgen Biotech Corp.), following the manufacturer’s protocol with minor modifications as detailed previously (37). Briefly, swabs were resuspended in PBS, pelleted, and subjected to bead-assisted thermal lysis. DNA was purified using column-based extraction with sequential buffer washes and eluted in 50 µl of elution buffer. DNA concentration and purity were assessed, and samples with an A260/A280 ratio of ∼1.8 were considered suitable for downstream analysis.

### 2.5 Amplification and 16S rRNA Gene Sequencing

The DNA extracted from the swabs was transported to the miBiome Therapeutics lab (Mumbai, India) for sequencing of the hypervariable V3-V4 region of the 16S rRNA gene. This region was amplified using universal barcoded primer pairs: 341F (5’-CCTACGGGNGGCWGCAG-3’) and 785R (5’-GACTACHVGGGTATCTAATCC-3’). Approximately 12.5 ng of swab DNA per sample underwent PCR amplification under optimized conditions, as previously describe^15^. The amplified products were purified and indexed using the Nextera XT Index Kit (Illumina). DNA library quantification was carried out according to lab-optimized protocols. The final libraries were pooled and sequenced on the Illumina MiSeq platform, utilizing v3

Reagent chemistry generating 2x300 bp paired-end reads.

### 2.6 Quality control and Sequence Processing for community structure analysis

Raw reads were assessed for quality using the FASTQC tool. The downstream analysis was conducted using four different pipelines to ensure robustness. In the first, the reads were merged using the FLASH assembler tool^25^. The assembled reads were analyzed for community structure using the QIIME pipeline (v2.0). Operational Taxonomic Unit (OTU) picking was performed with the UCLUST closed-reference method, and representative OTUs were assigned taxonomy using the RDP classifier method, with the GreenGenes (13_8 release) dataset as the reference. Data normalization was performed through the rarefaction step included in the QIIME package.

In the second pipeline, sequence analysis was also conducted using the 16S-based metagenomics workflow of MiSeq Reporter v2.3 (Illumina). In brief, sequences were demultiplexed based on index sequences to create FASTQ files. OTUs clustering and taxonomic classification were performed using the Illumina-curated version of the May 2013 release of the Greengenes database.

In the third and fourth method, analysis of sequenced data was done by using MOTHUR software (v.4.4) using two different databases. Data was analysed by following the standard operating procedure (SOP) from Schloss et al.(38) Reads were processed for OTU classification and taxonomic assignment using both Greengenes v13.8 and EzBioCloud databases (39,40).

### 2.7 Microbiome Diversity and Statistical Analysis

Microbial community structure and diversity were evaluated using multiple analytical approaches(39). Alpha diversity was measured using the Gini-Simpson index, while beta diversity was assessed through Bray–Curtis dissimilarity followed by Principal Coordinates Analysis (PCoA) to visualize compositional differences between samples. Beta diversity (Bray–Curtis): PERMANOVA was performed using the vegan package’s adonis function with the formula as Bray Distance ∼ Term where Bray Distance is the output of phyloseq’s distance function using the “bray” method on relative abundance data for species. The data was visualized using phyloseq’s plot_ordination function with the relative abundance data for species as input, method as “PCoA”, and distance as “bray”. The stat ellipse function of the ggplot2 package was used to draw ellipses for Vagitype assuming multivariate t-distribution showcasing Vagitypes that have enough points to calculate an ellipse. Differential abundance of microbial taxa was analyzed using unsupervised clustering. To explore patterns within the microbiome, samples were grouped into Community State Types (CSTs) based on dominant taxa profiles. All statistical analyses and visualisations were conducted using R (version 3.6.3), phyloseq version 1.28.0; vegan version 2.5-7; ggplot2 version 3.3.5; reshape2 version 1.4.4.

## 3. Results

### 3.1 Study Cohort and Sequencing

The analytical workflow is illustrated in Figure 1. After conducting initial quality assurance and control (QA/QC), removing chimeras, and aligning sequences, 371 operational taxonomic units (OTUs) were identified.

**Figure 1:**
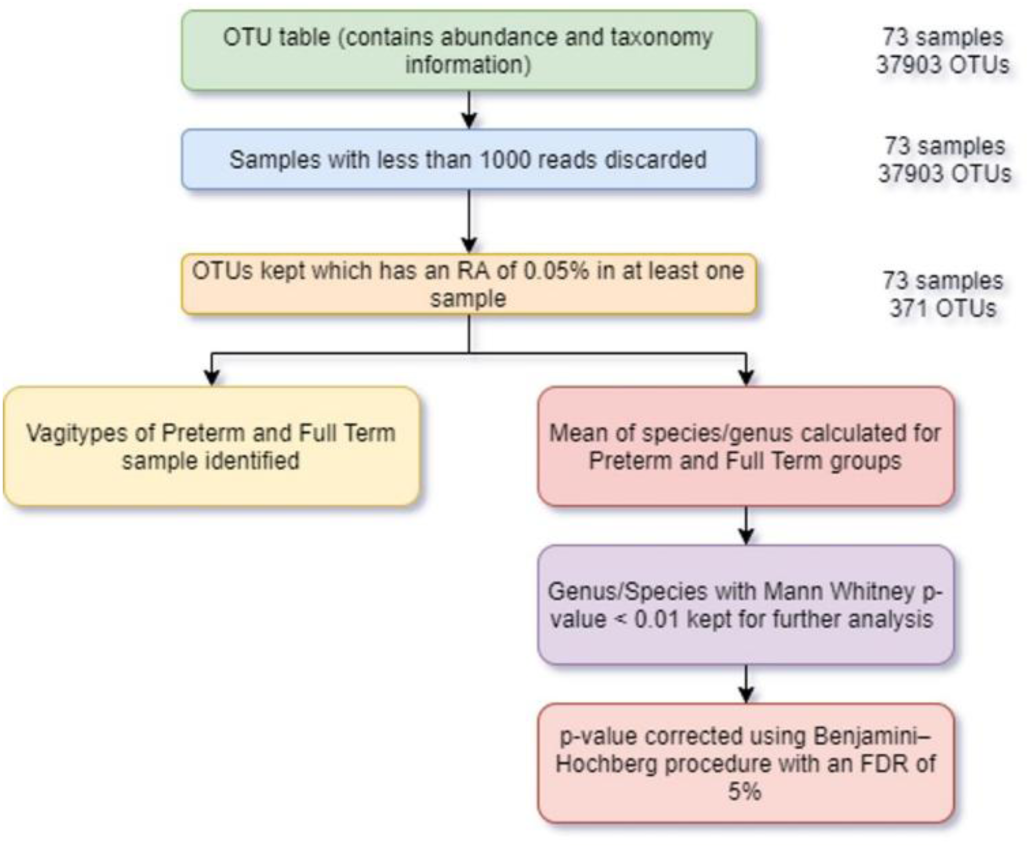
Data Analysis Workflow.

### 3.2 Microbial Diversity and Community Structure

At the time of delivery, Alpha diversity was comparable between women who delivered at term and those who delivered preterm (Fig. 2A, p = 0.24 Wilcoxon rank-sum test). The beta diversity was not significantly different between the two groups (Fig. 2B, R² value = 0.01878, p-value = 0.2146). Furthermore, Principal Coordinate Analysis (PCoA) revealed no distinct segregation between the two groups, indicating similar microbial compositions (Fig. 2C).

**Figure 2:**
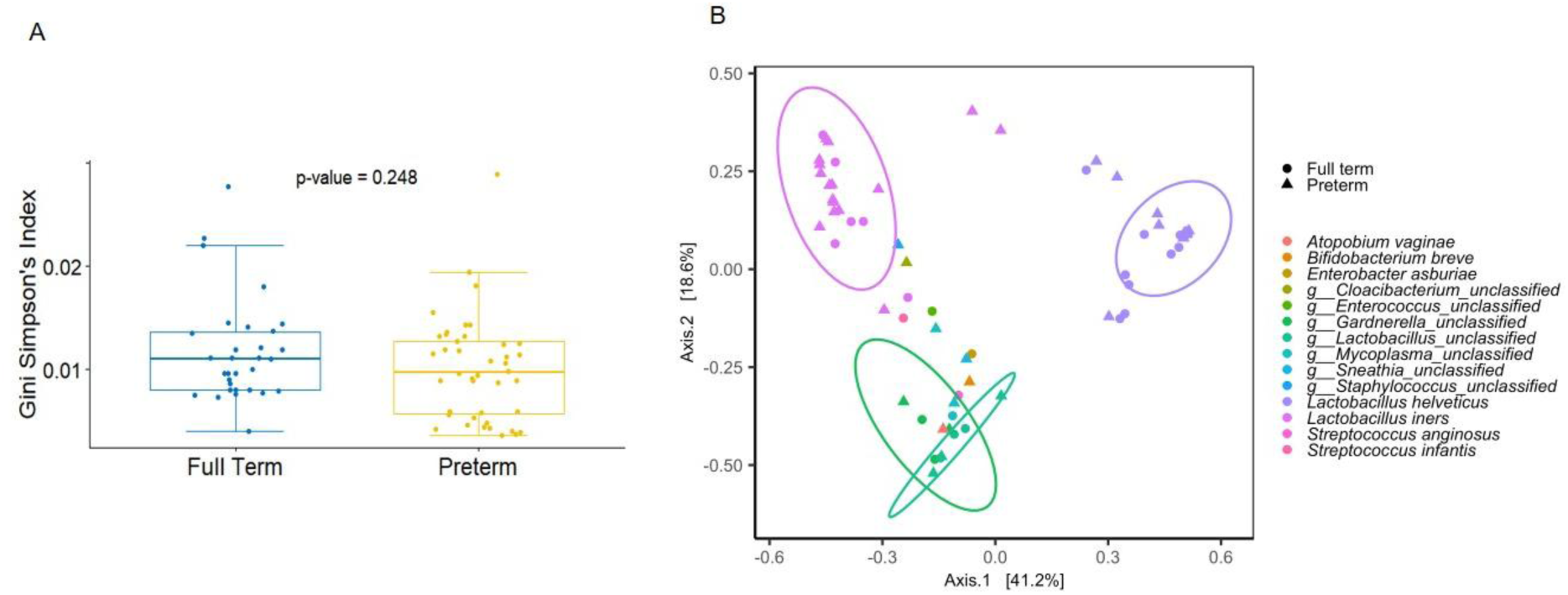
The alpha and beta diversity metrics of the vaginal microbiome were compared between preterm and term births. (A) Alpha (Gini Simpson) diversity between the two groups. (B) Beta diversity (Bray–Curtis): The colours correspond to the Vagitype for each sample i.e. assigned based on the most abundant bacteria in the vaginal microbiome of that sample, and listed in the legend. The sample term is showcased as a shape.

### 3.3 Vagitype Profile and Relative Abundance

At the genus level, *Lactobacillus* emerged as the most prevalent vagitype followed by *Streptococcus*, *Prevotella*, *Bifidobacterium*, and *Enterobacter* species. Analysis of the top 15 most abundant species revealed no significant differences in the relative abundance of these species between preterm and term births (Fig. 3). Given that different bioinformatics pipelines can influence the outcomes of microbiome analyses, we reanalysed the data using four distinct pipelines^28^ and compared the top 30–50 genera and species identified by these methods. Analyses were conducted using multiple bioinformatics pipelines, which yielded broadly consistent community-level trends.

**Figure 3:**
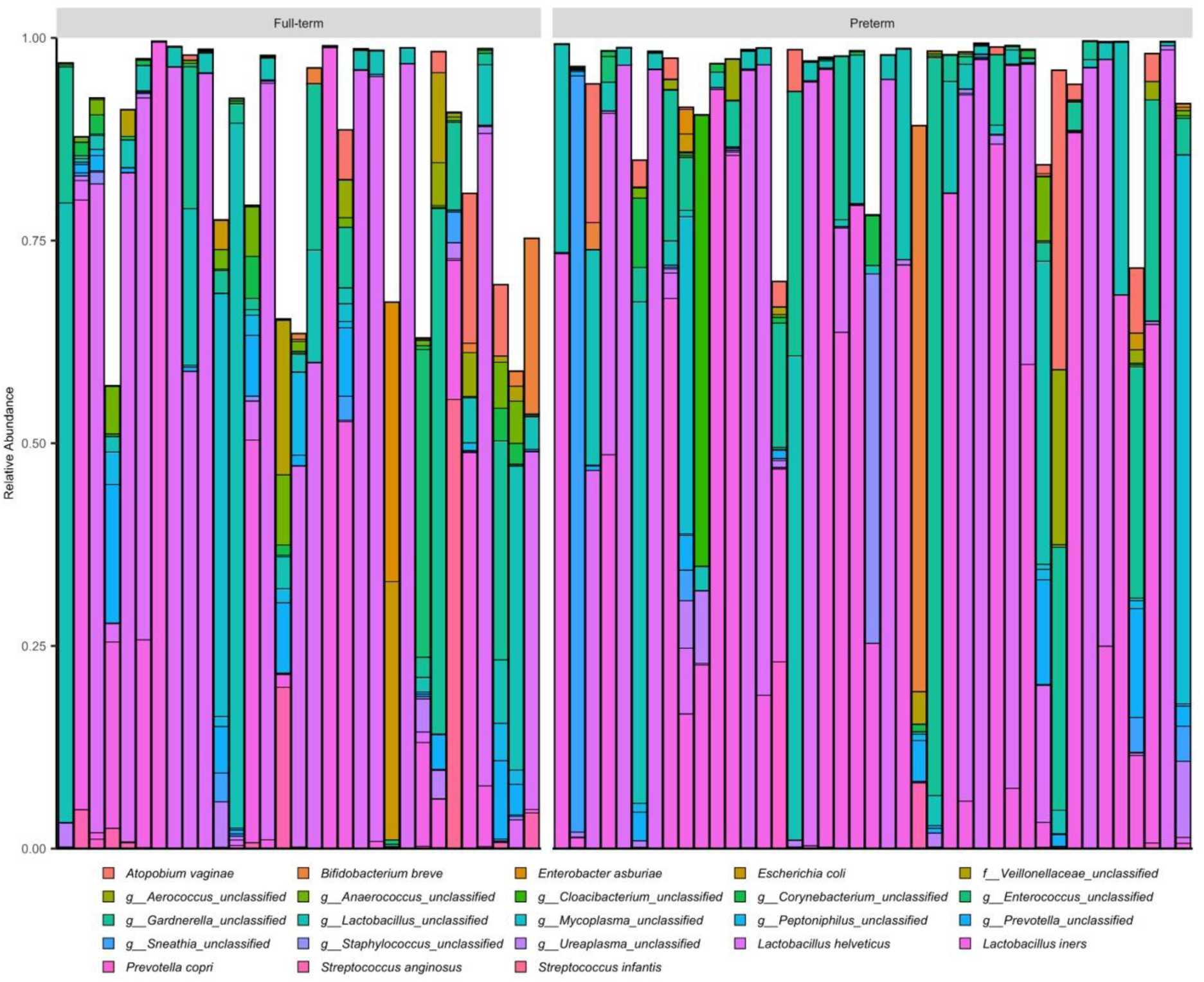
The relative abundance of reads assigned to the top 15 species-level taxa in matched preterm (n=41) and term (n=31) vaginal samples. The top 15 most abundant species for both groups and were determined using phyloseq’s taxa_sums function and plotted in a stacked barplot for each sample.

### 3.4 Clustering analysis

Supervised clustering of the microbial community data (Fig. 4) was unable to distinctly classify the samples into separate groups corresponding to preterm and term births. This lack of segregation further supports that there are no major differences in the vaginal microbiome composition between women delivering preterm and those delivering at term in the studied cohort.

**Figure 4:**
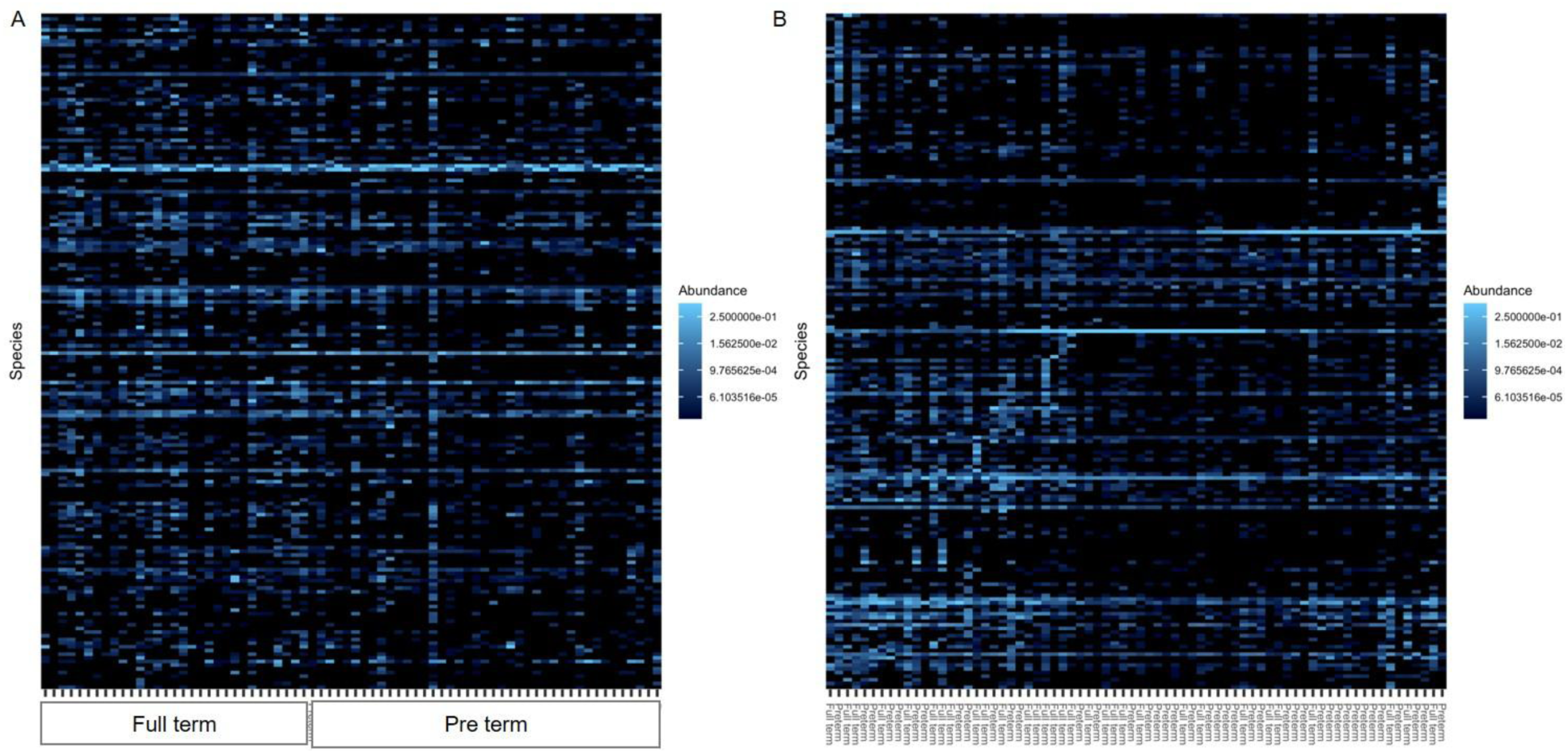
Clustering of the microbial communities in vaginal swabs of women delivering preterm (n=41) and term (n=31). (A) Heat map of unclustered samples classified based solely on the time of delivery. (B) Hierarchical clustering dendrograms (clustered by the PCoA) were generated using complete linkage. Each row represents a genus, and each column corresponds to a sample. The colour scale indicates the abundance of reads. Heatmap were generated using phyloseq package’s plot_heatmap function with ordination method as “PCoA” and distance as “bray”.

### 3.5 Lactobacillus Species Distribution in Preterm and Term Births

Our microbiome analysis identified several *Lactobacillus* species, with *L. helveticus* and *L. iners* being the most abundant across the samples (Fig.5). Additionally, *L. mucosae*, *L. hamsteri*, and *L. salivarius* were detected, though they were among the least common species in the group. The overall levels of *Lactobacillus* species were comparable between women who delivered preterm and those who delivered at term. No statistically significant differences were observed in the abundance of any individual *Lactobacillus* species between the two groups. While the mean abundance of *L. iners* was slightly higher in preterm birth samples compared to term birth samples, this difference was not statistically significant.

**Figure 5:**
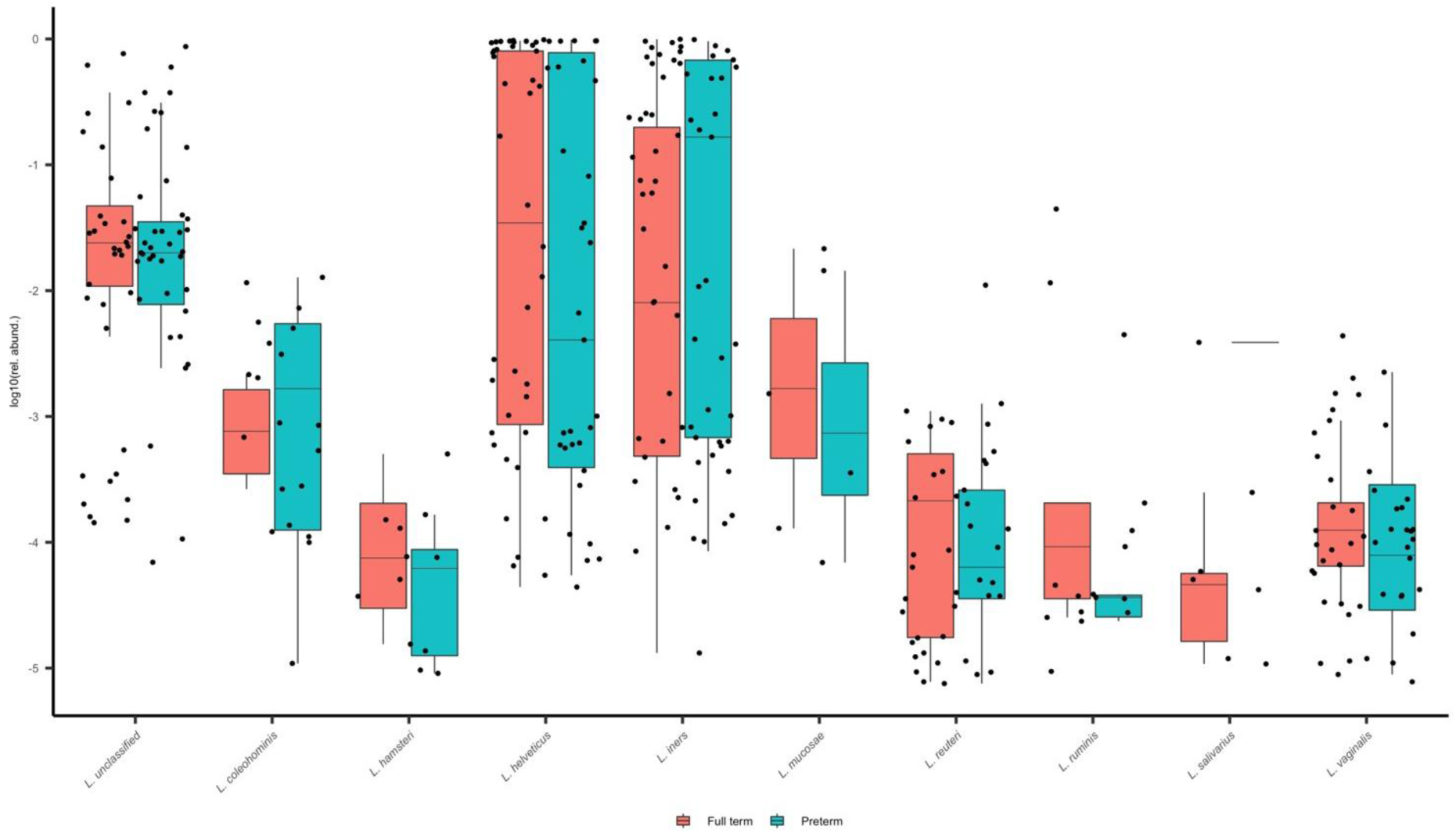
The abundance of various Lactobacillus species in vaginal swabs of women delivering preterm (n=41) and term (n=31). Differences in relative abundance of various Lactobacillus species between preterm and full-term groups were assessed using the Wilcoxon rank-sum test. P-values were adjusted for multiple comparisons using the Benjamini–Hochberg false discovery rate, Boxplots display log10-scaled relative abundances for visualization.

### 3.6 Non-Lactobacillus Taxa in the Context of Preterm Birth

When examining the relative abundance of non-*Lactobacillus* bacterial taxa that may be associated with preterm births (Fig. 6), *Peptostreptococcus* and *Streptococcus* genera were significantly more abundant in women who delivered preterm compared to those who delivered at term among the top 18 genera identified in the vaginal swabs. However, after adjustment only the levels of Streptococcus genus was significantly different between the two groups (P-value = 0.032 Wilcoxon rank-sum test). The species level, post adjustments, there were no significant differences in any of the organisms in women with preterm as compared to term delivery.

**Figure 6:**
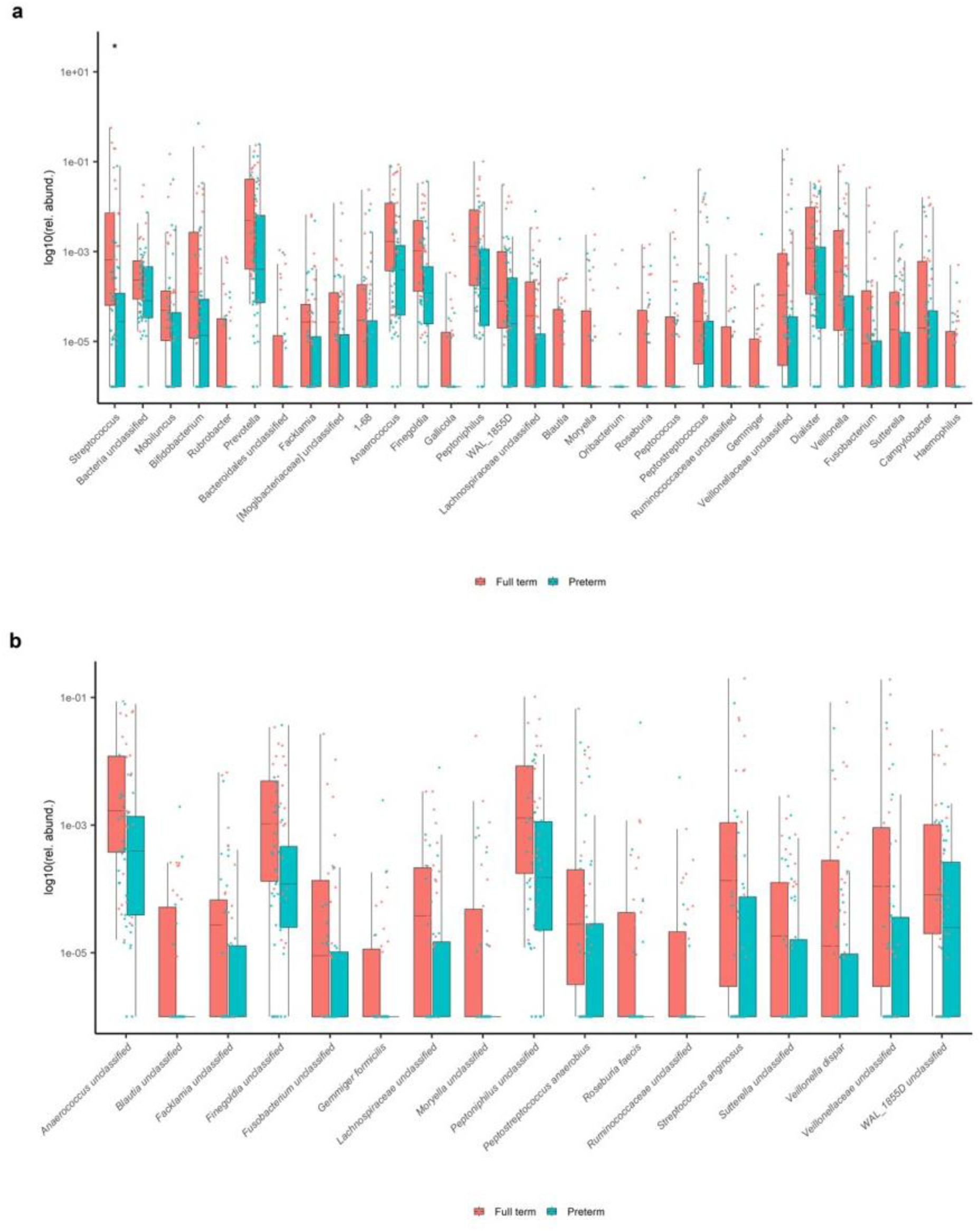
The abundance of selected bacterial (a) genus and (b) species, in vaginal swabs of women delivering preterm (n=41) and term (n-31). Differences in relative abundance between preterm and full-term groups were assessed using the Wilcoxon rank-sum test. P-values were adjusted for multiple comparisons using the Benjamini–Hochberg false discovery rate, Boxplots display log10-scaled relative abundances for visualization. * values significantly different between the two groups (p=0.032).

## 4. Discussion

Preterm birth (PTB) remains a leading global cause of neonatal morbidity and mortality, with India contributing over one-quarter of the worldwide PTB burden. Identifying modifiable risk factors is critical to reducing their prevalence. The vaginal microbiome plays a crucial role in women’s reproductive health, affecting fertility, pregnancy, and infection susceptibility. A healthy vaginal microbiome is often dominated by *Lactobacillus* species, which help maintain an acidic environment that suppresses pathogenic colonization. Disruptions in the vaginal microbiome can lead to complications, including preterm birth. Traditional culture-based methods and nucleic acid analysis have implicated several bacterial species, including *Group B Streptococcus* (GBS), *Escherichia coli*, and *Gardnerella vaginalis* in preterm birth, which can alter the vaginal microbiome and contribute to early delivery (17,36,41).

Recent advancements in high-throughput 16S rRNA sequencing have expanded our understanding by enabling analysis of a broader range of uncultivable and low-abundance bacterial species. Numerous studies across different populations have aimed to identify specific vagitypes that may contribute to preterm births, with inconsistent findings across populations (24,31–36,42–44). Factors such as host genetics, vaginal hygiene practices, and antibiotic usage are thought to contribute to these discrepancies. Additionally, ethnic variation in vaginal microbiota diversity has also been reported. Meta-analysis of global data revealed Black women tend to harbour a lower abundance of Lactobacillus and higher abundance of pathogenic microbes like *Megasphaera dialister*, *Staphylococcus*, and *Aerococcus* (31,45).

Despite India’s high PTB burden, there remains a limited number of studies on the vaginal microbiome of Indian women (34–36). In our cross-sectional case-control study (conducted at the time of delivery) from western India, no significant differences were observed in either alpha or beta diversity between the vaginal microbiota of women who delivered preterm or at term, suggesting no significant vaginal dysbiosis associated with PTB in our cohort. These results aligns with findings from some studies that found no major association of vaginal microbiome and PTB (29). Assessment of the relative distribution of the top 15 species between women who delivered preterm and those who delivered at term, and unsupervised clustering could not differentiate preterm and term births, reinforcing the absence of distinct microbial signatures for preterm and term births classification based on the vaginal microbiome.

In contrast to our findings, longitudinal analyses from the GARBH-Ini cohort from North India demonstrated enrichment of *Lactobacillus iners* and selected anaerobic taxa in women who delivered preterm; however, the largest differences were observed in the first trimester, with attenuation of these differences by the time of delivery (27). More recently, an expanded GARBH-Ini study using metagenomic approaches (35), as well as an independent cohort from western India (36), further confirmed that early pregnancy microbial signatures and community state types (CSTs) are predictive of PTB. In contrast, our cross-sectional analysis at the time of delivery revealed no major differences in overall microbial diversity or community structure between term and preterm births. This discrepancy likely reflects differences in sampling time points, as our samples were obtained at delivery, in contrast to studies capturing vaginal microbiome dynamics across pregnancy trimesters. Taken together, these findings suggest that microbiome-associated risk for preterm birth in Indian populations is established earlier in gestation, and that vaginal microbial profiles at delivery may represent a convergent or stabilized state rather than etiological processes driving preterm labour.

Interestingly, we identified a modest but statistically significant increase in the relative abundance of *Streptococcus* in the preterm group. While some members of the *Streptococcus* family are associated with reproductive tract infections and inflammation leading to PTB (17,45), given the limited effect size and the lack of broader microbial shifts, these findings may represent secondary changes rather than causal contributors. Longitudinal sampling earlier in gestation may help clarify whether these taxa precede or follow inflammatory or physiological changes that lead to preterm labour. Further studies should analyse quantitative differences in microbial load and validate these findings in larger cohorts.

Our study has several strengths, including rigorous sample inclusion criteria, robust quality control, and the use of multiple analytical platforms to confirm findings. However, the cross-sectional design, with sampling done at the time of delivery, may not capture microbial alterations occurring earlier in pregnancy when the risk for PTB may be biologically imprinted. Also, a modest sample size may limit the ability to detect subtle differences in less abundant taxa or microbiome functions.

In conclusion, in this cross-sectional cohort of samples collected at time of delivery, we did not find major differences in microbial diversity or composition between term and preterm deliveries, suggesting that vaginal dysbiosis at delivery may not be a key driver of PTB in this population. Larger, longitudinal studies that capture microbial dynamics across pregnancy, coupled with functional and host-response analyses, are essential to better understand the role of the vaginal microbiome in preterm birth risk and to inform targeted prevention strategies in high-burden settings.

## Authors Contributions

AS lead the study, performed all the microbiological and molecular assays & data analysis. AH, NV, SK optimized the molecular assay and assisted in performing the same. SS collected the swabs samples, administered the PIS and got consent. GD, GS, SK, AR, CA, AK were involved in DNA sequencing, NGS data generation and data interpretation. SK coordinated the clinical arm of the study, contributed to the sample collection and patient selection. ADU and DM conceptualized the study. AS and DM analysed the data and conceptualized the manuscript. GS and KC contributed to data analysis, wrote the manuscript, and edited the draft. All the authors finalized the manuscript and approved the same.

## Data Availability

Data produce in the study are contained in the manuscript

## Acknowledgment

ChatGPT5.2 was used for language editing with human supervision. The study was supported by MGMIHS (SEED grant). The DM lab at ICMR-NIRRCH is supported by grants from Indian Council for Medical Research. KC and NV acknowledge the Fulbright-Nehru Fellowship. AH acknowledges ICMR postdoctoral fellowship.

## References

1. Ohuma EO, Moller AB, Bradley E, Chakwera S, Hussain-Alkhateeb L, Lewin A, et al. National, regional, and global estimates of preterm birth in 2020, with trends from 2010: a systematic analysis. The Lancet. 2023 Oct 7;402(10409):1261–71. doi:10.1016/S0140-6736(23)00878-4 PubMed PMID: 37805217.

2. Dixit P, Bhatia M, Bhatia C, Dwivedi LK, Singh S. Risk factors for preterm birth and its effect on neonatal mortality in India: evidence from the National Health Family Survey-5. Discover Public Health 2025 22:1. 2025 Aug 18;22(1):481-. doi:10.1186/s12982-025-00868-0

3. Jana A. Correlates of low birth weight and preterm birth in India. PLOS ONE. 2023 Aug 17;18(8):e0287919. doi:10.1371/journal.pone.0287919 PubMed PMID: 37590211.

4. Baud A, Hillion KH, Plainvert C, Tessier V, Tazi A, Mandelbrot L, et al. Microbial diversity in the vaginal microbiota and its link to pregnancy outcomes. Scientific Reports 2023 13:1. 2023 Jun 4;13(1):9061-. doi:10.1038/s41598-023-36126-z PubMed PMID: 37271782.

5. Young RB, Correia GDS, MacIntyre DA. Host, microbial and environmental drivers of vaginal microbiota composition. Fertility and Sterility. 2026 Jan 27. doi:10.1016/j.fertnstert.2026.01.020

6. Goswami A, Ghosh S, Bandyopadhyay A, Saha RG, Sengupta P, Bhuniya U, et al. Comparative Analysis of Vaginal Microbiome Associated with Oncogenic HPV Infection Among Different Ethnic Groups of Women of the Eastern Region of India. Indian journal of microbiology. 2025 Dec 1;65(4):1877–90. doi:10.1007/s12088-024-01320-8 PubMed PMID: 41424906.

7. Tandon D, Shah N, Goriwale M, Karandikar K, Begum S, Patil AD, et al. Mapping the vaginal microbiota variations in women from a community clinic in Mumbai, India. Indian Journal of Medical Microbiology. 2023 Sep 1;45. doi:10.1016/j.ijmmb.2023.100393 PubMed PMID: 37573043.

8. Xie Z, Chen Z, Ma G. Dynamic changes in the pregnancy microbiome and their role in preterm birth. Frontiers in cellular and infection microbiology. 2025;15. doi:10.3389/fcimb.2025.1683610 PubMed PMID: 41488481.

9. Pramanick R, Nathani N, Warke H, Mayadeo N, Aranha C. Vaginal Dysbiotic Microbiome in Women With No Symptoms of Genital Infections. Frontiers in Cellular and Infection Microbiology. 2022 Jan 12;11. doi:10.3389/fcimb.2021.760459 PubMed PMID: 35096634.

10. Avitabile E, Menotti L, Croatti V, Giordani B, Parolin C, Vitali B. Protective Mechanisms of Vaginal Lactobacilli against Sexually Transmitted Viral Infections. International Journal of Molecular Sciences 2024, Vol 25, Page 9168. 2024 Aug 23;25(17):9168. doi:10.3390/ijms25179168 PubMed PMID: 39273118.

11. Pagar R, Deshkar S, Mahore J, Patole V, Deshpande H, Gandham N, et al. The microbial revolution: Unveiling the benefits of vaginal probiotics and prebiotics. Microbiological Research. 2024 Sep 1;286. doi:10.1016/j.micres.2024.127787 PubMed PMID: 38851010.

12. Pawar K, Aranha C. Lactobacilli metabolites restore E-cadherin and suppress MMP9 in cervical cancer cells. Current Research in Toxicology. 2022 Jan 1;3. doi:10.1016/j.crtox.2022.100088

13. Valenti P, Rosa L, Capobianco D, Lepanto MS, Schiavi E, Cutone A, et al. Role of lactobacilli and lactoferrin in the mucosal cervicovaginal defense. Frontiers in Immunology. 2018 Mar 1;9(MAR):338405. doi:10.3389/fimmu.2018.00376

14. Nair VG, Chellappan DR, Durai RD, Y.B.R.D R, Narbhavi D, A. A, et al. Synergistic antipersister, efflux inhibitory & antibiofilm activities of vaginal Lactobacillus-derived postbiotics against UPEC: toward a novel therapeutic for utis. Scientific reports. 2026 Jan 10;16(1). doi:10.1038/s41598-026-35736-7 PubMed PMID: 41520010.

15. Surve MV, Anil A, Kamath KG, Bhutda S, Sthanam LK, Pradhan A, et al. Membrane Vesicles of Group B Streptococcus Disrupt Feto-Maternal Barrier Leading to Preterm Birth. PLOS Pathogens. 2016 Sep 1;12(9):e1005816. doi:10.1371/journal.ppat.1005816 PubMed PMID: 27583406.

16. Kurian NK, Modi D. Mechanisms of group B Streptococcus-mediated preterm birth: lessons learnt from animal models. Reproduction & fertility. 2022 Jul 1;3(3):R109–20. doi:10.1530/RAF-21-0105 PubMed PMID: 35794927.

17. Ashary N, Singh A, Chhabria K, Modi D. Meta-analysis on prevalence of vaginal group B streptococcus colonization and preterm births in India. Journal of Maternal-Fetal and Neonatal Medicine. 2022;35(15):2923–31. doi:10.1080/14767058.2020.1813705 PubMed PMID: 32873095.

18. Tellapragada C, Eshwara VK, Bhat P, Kamath A, Aletty S, Mukhopadhyay C. Screening of vulvovaginal infections during pregnancy in resource constrained settings: Implications on preterm delivery. Journal of Infection and Public Health. 2017 Jul 1;10(4):431–7. doi:10.1016/j.jiph.2016.06.003 PubMed PMID: 27422139.

19. Payne MS, Newnham JP, Doherty DA, Furfaro LL, Pendal NL, Loh DE, et al. A specific bacterial DNA signature in the vagina of Australian women in midpregnancy predicts high risk of spontaneous preterm birth (the Predict1000 study). American Journal of Obstetrics and Gynecology. 2021 Feb 1;224(2):206.e1-206.e23. doi:10.1016/j.ajog.2020.08.034 PubMed PMID: 32861687.

20. Kandari S, Kandari S. A Review on the Role of Endometrial Microbiome in Reproductive Pathologies Affecting Female Infertility. Fertility Science and Research. 2024 Jun 15;11(5):5. doi:10.25259/fsr_43_23

21. Patel N, Patel N, Pal S, Nathani N, Pandit R, Patel M, et al. Distinct gut and vaginal microbiota profile in women with recurrent implantation failure and unexplained infertility. BMC women’s health. 2022 Dec 1;22(1). doi:10.1186/s12905-022-01681-6 PubMed PMID: 35413875.

22. Chopra C, Kumar V, Kumar M, Bhushan I. Role of vaginal microbiota in idiopathic infertility: a prospective study. Microbes and Infection. 2024 May 1;26(4). doi:10.1016/j.micinf.2024.105308 PubMed PMID: 38311069.

23. Holliday M, Uddipto K, Castillo G, Vera LE, Quinlivan JA, Mendz GL. Insights into the Genital Microbiota of Women Who Experienced Fetal Death in Utero. Microorganisms. 2023 Aug 1;11(8). doi:10.3390/microorganisms11081877 PubMed PMID: 37630436.

24. Kulshrestha S, Narad P, Singh B, Pai SS, Vijayaraghavan P, Tandon A, et al. Biomarker Identification for Preterm Birth Susceptibility: Vaginal Microbiome Meta-Analysis Using Systems Biology and Machine Learning Approaches. American journal of reproductive immunology (New York, NY : 1989). 2024 Jul 1;92(1). doi:10.1111/aji.13905 PubMed PMID: 39033501.

25. Feehily C, Crosby D, Walsh CJ, Lawton EM, Higgins S, McAuliffe FM, et al. Shotgun sequencing of the vaginal microbiome reveals both a species and functional potential signature of preterm birth. NPJ biofilms and microbiomes. 2020 Dec 1;6(1). doi:10.1038/s41522-020-00162-8 PubMed PMID: 33184260.

26. Liao J, Shenhav L, Urban JA, Serrano M, Zhu B, Buck GA, et al. Microdiversity of the vaginal microbiome is associated with preterm birth. Nature communications. 2023 Dec 1;14(1). doi:10.1038/s41467-023-40719-7 PubMed PMID: 37591872.

27. Kumar S, Kumari N, Talukdar D, Kothidar A, Sarkar M, Mehta O, et al. The Vaginal Microbial Signatures of Preterm Birth Delivery in Indian Women. Frontiers in Cellular and Infection Microbiology. 2021 May 13;11. doi:10.3389/fcimb.2021.622474 PubMed PMID: 34094994.

28. Romero R, Hassan SS, Gajer P, Tarca AL, Fadrosh DW, Bieda J, et al. The vaginal microbiota of pregnant women who subsequently have spontaneous preterm labor and delivery and those with a normal delivery at term. Microbiome. 2014 May 27;2(1):18. doi:10.1186/2049-2618-2-18 PubMed PMID: 24987521.

29. Gulavi E, Mwendwa F, Atandi DO, Okiro PO, Hall M, Beiko RG, et al. Vaginal microbiota in women with spontaneous preterm labor versus those with term labor in Kenya: a case control study. BMC Microbiology 2022 22:1. 2022 Nov 10;22(1):270-. doi:10.1186/s12866-022-02681-0 PubMed PMID: 36357861.

30. Kulshreshtha S, Narad P, Singh B, Modi D, Sengupta A. Identification of Distinct Vaginal Microbiota Signatures Contributing Toward Preterm Birth Using an Integrative Computational Approach. Microbiology and Biotechnology Letters. 2023 Mar 1;51(1):109–23. doi:10.48022/mbl.2210.10008

31. Juliana NCA, Suiters MJM, Al-Nasiry S, Morré SA, Peters RPH, Ambrosino E. The Association Between Vaginal Microbiota Dysbiosis, Bacterial Vaginosis, and Aerobic Vaginitis, and Adverse Pregnancy Outcomes of Women Living in Sub-Saharan Africa: A Systematic Review. Frontiers in Public Health. 2020 Dec 10;8:567885. doi:10.3389/fpubh.2020.567885 PubMed PMID: 33363078.

32. Callahan BJ, DiGiulio DB, Aliaga Goltsman DS, Sun CL, Costello EK, Jeganathan P, et al. Replication and refinement of a vaginal microbial signature of preterm birth in two racially distinct cohorts of US women. Proceedings of the National Academy of Sciences of the United States of America. 2017 Sep 12;114(37):9966–71. doi:10.1073/pnas.1705899114 PubMed PMID: 28847941.

33. Wang L, Zhang J, Zhang M, Xu Z, Zheng Y, Lv B, et al. Insights into the alteration of vaginal microbiota and metabolites in pregnant woman with preterm delivery: prospective cohort study. Frontiers in cellular and infection microbiology. 2025;15. doi:10.3389/fcimb.2025.1580801 PubMed PMID: 41050755.

34. Talukdar D, Raju YJ, Jana P, Sharma K, Babele P, Kothidar A, et al. Genomic insights into the potency and functional roles of Lactobacillus species in term and preterm births. Genomics. 2025 Jul 1;117(4). doi:10.1016/j.ygeno.2025.111063 PubMed PMID: 40456421.

35. Talukdar D, Sarkar M, Ahrodia T, Kumar S, De D, Nath S, et al. Previse preterm birth in early pregnancy through vaginal microbiome signatures using metagenomics and dipstick assays. iScience. 2024 Nov 15;27(11). doi:10.1016/j.isci.2024.111238

36. Patel DC, Rao A, Patel VB, Mistry TU. Impact of Early Pregnancy Vaginal Microbiome Composition on Preterm Birth Risk: A Prospective Cohort Study. European Journal of Cardiovascular Medicine. 2025 May 15;15:283–6. doi:10.61336/EJCM/25-05-51

37. Colaco S, Chhabria K, Singh D, Bhide A, Singh N, Singh A, et al. Expression map of entry receptors and infectivity factors for pan-coronaviruses in preimplantation and implantation stage human embryos. Journal of Assisted Reproduction and Genetics. 2021 Jul 1;38(7):1709–20. doi:10.1007/S10815-021-02192-3, PubMed PMID: 33913101.

38. Schloss PD, Westcott SL, Ryabin T, Hall JR, Hartmann M, Hollister EB, et al. Introducing mothur: Open-source, platform-independent, community-supported software for describing and comparing microbial communities. Applied and Environmental Microbiology. 2009 Dec;75(23):7537–41. doi:10.1128/AEM.01541-09 PubMed PMID: 19801464.

39. Bramble MS, Vashist N, Ko A, Priya S, Musasa C, Mathieu A, et al. The gut microbiome in konzo. Nature Communications 2021 12:1. 2021 Sep 10;12(1):5371-. doi:10.1038/s41467-021-25694-1 PubMed PMID: 34508085.

40. Chalita M, Kim YO, Park S, Oh HS, Cho JH, Moon J, et al. EzBioCloud: a genome-driven database and platform for microbiome identification and discovery. International Journal of Systematic and Evolutionary Microbiology. 2024 Jun 18;74(6):006421. doi:10.1099/ijsem.0.006421 PubMed PMID: 38888585.

41. Sirisha P, Prashanth S, Arun P, Barathi A. Association Between Spontaneous Preterm Labor and Genitourinary Tract Infections Among Pregnant Women in a Tertiary Care Hospital in South India: A Cross-Sectional Study. Cureus. 2024 Oct 21;16(10). doi:10.7759/cureus.71973 PubMed PMID: 39569252.

42. Wang L, Zhang J, Zhang M, Xu Z, Zheng Y, Lv B, et al. Insights into the alteration of vaginal microbiota and metabolites in pregnant woman with preterm delivery: prospective cohort study. Frontiers in cellular and infection microbiology. 2025;15. doi:10.3389/fcimb.2025.1580801 PubMed PMID: 41050755.

43. Stout MJ, Zhou Y, Wylie KM, Tarr PI, Macones GA, Tuuli MG. Early pregnancy vaginal microbiome trends and preterm birth. American Journal of Obstetrics and Gynecology. 2017 Sep 1;217(3):356.e1-356.e18. doi:10.1016/j.ajog.2017.05.030 PubMed PMID: 28549981.

44. Huang C, Gin C, Fettweis J, Foxman B, Gelaye B, MacIntyre DA, et al. Meta-analysis reveals the vaginal microbiome is a better predictor of earlier than later preterm birth. BMC biology. 2023 Dec 1;21(1). doi:10.1186/s12915-023-01702-2 PubMed PMID: 37743497.

45. Kulshrestha S, Narad P, Pai SS, Singh B, Modi D, Sengupta A. Metagenomic investigation of 16S rRNA marker gene samples to analyze the role of race, ethnicity, and location in preterm birth: A comprehensive vaginal microbiome meta-analysis. Human Gene. 2024 Feb 1;39:201260. doi:10.1016/j.humgen.2024.201260

